# Effects of estradiol on biological age measured using the glycan age index

**DOI:** 10.1101/2020.06.25.20138503

**Authors:** Julija Jurić, Wendy M. Kohrt, Domagoj Kifer, Marija Pezer, Peter A. Nigrovic, Gordan Lauc

**Author notes:** Corresponding address: Department of Biochemistry and Molecular Biology, Faculty of Pharmacy and Biochemistry, University of Zagreb, A. Kovačića 1, 10000 Zagreb, Croatia. E-mail addresses.

## Abstract

Glycan age is a recently developed biomarker based on glycans attached to immunoglobulin G (IgG). In large population cohorts glycan age associates well with lifestyle and disease-risk biomarkers, while some studies suggested that change in glycans precede development of several age-associated diseases. In this study we evaluated effects of estrogen on the glycan age. Gonadal hormones were suppressed in 36 healthy young women by gonadotropin releasing hormone agonist therapy for 6 months. In 15 of them estradiol was supplemented, while 21 received placebo resulting in very low estrogen levels during intervention. IgG was isolated from plasma samples before intervention, after 6 months of intervention and after subsequent 4-month recovery. In the placebo group the removal of gonadal hormones resulted in median increase of glycan age for 9.1 years (IQR 6.8 – 11.5 years, p = 3.73×10^−8^), which was completely prevented by transdermal estradiol supplementation. After the recovery period glycan age returned to baseline values also in the placebo group. These results suggest that IgG glycans and consequently also the glycan age are under strong influence of gonadal hormones and that hormone replacement therapy can prevent the increase of glycan age that occurs in the perimenopausal period.

## Introduction

The existence of inter-individual differences in the pace of biological ageing is an intriguing concept that tries to explain why some people stay healthy until very late chronological age, while other people age faster and have a shorter life expectancy. A number of biomarkers aimed at an objective estimation of biological age have been developed in the past several years, one of them being the glycan age, which is based on analysing glycans attached to immunoglobulin G (IgG) [1]. A key feature of a good biomarker of biological age is that the difference between chronological and biological age should correlate with known biomarkers of an unhealthy lifestyle and that increased biological age should predict future disease development. Glycans attached to IgG change significantly with age [1] and have been suggested as a promising biomarker of biological age[2]. Furthermore, since glycosylation affects interactions between IgG and different Fcγ receptors and other ligands, changes in glycosylation have direct effects on the function of the immune system[3], with multiple functional implications. IgG glycosylation is altered in many diseases[4] and in some cases these changes were found to occur before the onset of other symptoms [5–7].

Large population studies [1,8] and our recent study of intervention cohort suggest that estrogen regulates IgG glycosylation [9], which may explain why IgG glycome in premenopausal females reflects apparent lower biological age. Unfortunately, the published estrogen intervention study was based on the analysis of glycans released from the total plasma proteome and this prevented the calculation of glycan age from the available data since glycan age is based on IgG glycans. Aiming to evaluate the effects of ovarian sex hormones suppression followed by estradiol supplementation on biological age measured by the glycan age we reanalysed samples from the same intervention study using state of the art glycoprofiling technology [10].

## Results

IgG glycosylation was analysed in 36 healthy premenopausal women that were treated on an investigational basis with the gonadotropin-releasing hormone (GnRH) analogue leuprolide to lower gonadal steroids to postmenopausal levels and then randomized to placebo or transdermal estradiol (Figure 1) [11]. Plasma samples were collected at baseline (T1), after five months of hormonal suppression by monthly leuprolide injections plus either estrogen or placebo patches (T2), and four months after the end of intervention when natural hormonal cycling was restored (T3). The concentration of hormones at the baseline and differences from the baseline after intervention and after recovery timepoint are presented in Table 1.

**Table 1.** The concentration of hormones at baseline and differences from baseline after the intervention and at recovery.

| VARIABLE | INTERVENTION | CONCENTRATION<br>AT BASELINE<br>median (IQR) | DIFFERENCE IN CONCENTRATION RELATIVE TO BASELINE |  |  |  |
| --- | --- | --- | --- | --- | --- | --- |
| | | | Intervention<br>median (IQR) | $p_i$ | Recovery<br>median (IQR) | $p_R$ |
| **estradiol**<br>*pg/mL* | Placebo | 54.0<br>(44.5 - 79.2) | -31.5<br>(-53.5 to -19.8) | **0,001** | -7.5<br>(-45.2 to 25.5) | 0,989 |
|  | Estradiol | 57<br>(46 - 78) | -15.0<br>(-35.0 to 29.5) |  | -1<br>(-32 to 30) |  |
| **estrone**<br>*pg/mL* | Placebo | 52<br>(37 - 67) | -19<br>(-32 to -11) | **0,001** | -1<br>(-16 to 13) | 0,989 |
|  | Estradiol | 55.0<br>(39.0 - 68.0) | 3.0<br>(-10.5 to 20.0) |  | 0.0<br>(-10.5 to 23.0) |  |
| **FSH**<br>*mIU/mL* | Placebo | 5.95<br>(4.40 - 8.02) | -1.10<br>(-3.35 to -0.05) | **0,001** | 0.00<br>(-1.15 to 1.40) | 0,471 |
|  | Estradiol | 6.60<br>(4.95 - 9.15) | -5.10<br>(-7.25 to -3.05) |  | -1.85<br>(-3.03 to 0.45) |  |
| **LH**<br>*mIU/mL* | Placebo | 4.60<br>(3.55 - 5.10) | -4.00<br>(-4.88 to -2.88) | 0,768 | -0.700<br>(-1.450 to 0.725) | 0,815 |
|  | Estradiol | 4.90<br>(3.00 - 6.55) | -4.70<br>(-6.05 to -2.70) |  | -0.70<br>(-3.35 to 0.35) |  |
| **progesterone**<br>*ng/mL* | Placebo | 0.4<br>(0.3 - 0.7) | -0.1<br>(-0.4 to 0.0) | 0,989 | 0.0<br>(-0.2 to 0.2) | 0,760 |
|  | Estradiol | 0.4<br>(0.3 - 0.6) | -0.1<br>(-0.2 to 0.0) |  | 0.1<br>(-0.2 to 0.6) |  |
| **SHBG**<br>*nmol/L* | Placebo | 52<br>(30 - 63) | -8<br>(-16 to -2) | 0,094 | 2<br>(-6 to 5) | 0,760 |
|  | Estradiol | 35.0<br>(29.5 - 54.0) | -2.0<br>(-6.5 to 8.5) |  | -1.0<br>(-5.0 to 1.5) |  |
| **testosterone**<br>*ng/dL* | Placebo | 28.0<br>(17.0 - 32.2) | -4.0<br>(-10.8 to 0.0) | 0,760 | 0.0<br>(-2.0 to 6.9) | 0,760 |
|  | Estradiol | 29.0<br>(23.5 - 35.5) | -4.0<br>(-8.5 to 1.0) |  | 0.0<br>(-3.0 to 7.0) |  |

p values describe statistical significance of difference between estradiol and placebo group after intervention ( $p_i$ ) and recovery ( $p_R$ ). p values smaller than 0.05 in bold.
IQR – limits of the interquartile range (1<sup>st</sup> quartile - 3<sup>rd</sup> quartile).
FSH - Follicle-stimulating hormone; LH - Luteinizing hormone; SHBG - sex hormone-binding globulin

**Figure 1.**
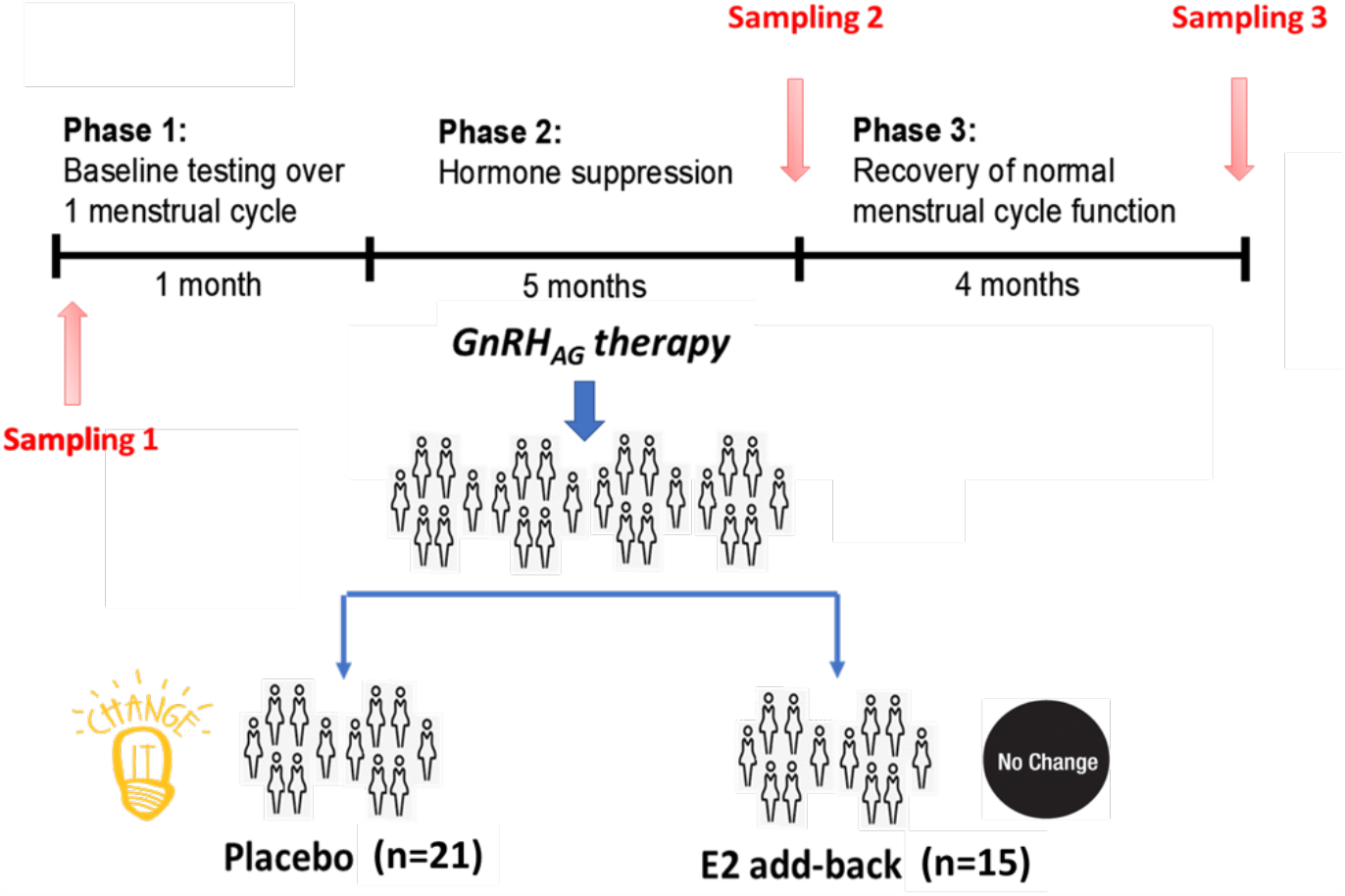
Design of the gonadal hormone suppression intervention study.

Suppression of ovarian sex hormones production resulted in a median increase of glycan age by 9.1 years, which was completely abolished by estradiol supplementation (Figure 2, Table 2). Both the extent of change in hormone levels (Table 1) and the extent of change in glycan age varied significantly, thus we wondered whether the estradiol baseline levels or the extent of changes in estradiol levels correlated with the extent of change in glycan age. The analysis did not reveal any statistically significant correlation between these two parameters (Figures 3A and 3B). Then we checked whether the change in glycan age correlated with baseline chronological age, baseline glycan age or the difference between chronological and glycan ages. Intuitively one would expect a larger increase in glycan age in chronologically younger women and indeed we did observe negative correlation between the extent of change induced by suppression of gonadal hormones and age (r = - 0.54, p = 1.1e-02, Figure 3E). Interestingly, much stronger correlation was observed for the initial glycan age (r = −0.84, p = 1.57e-06, Figure 3C) and the difference between glycan age and the chronological age (r = −0,66, p = 1,07E-03, Figure 3D) suggesting that low glycan age is strongly dependant on gonadal hormones.

**Table 2.** Chronological age (years) at the baseline, and differences in glycan age relative to the baseline after the intervention and after recovery timepoint.

| INTERVENTION | AGE (YEARS)<br>AT BASELINE<br>median (IQR) | DIFFERENCE IN GLYCANAGE (YEARS)<br>RELATIVE TO BASELINE. SAMPLING AFTER: |  |  |  |
| --- | --- | --- | --- | --- | --- |
|  |  | Intervention<br>median (IQR) | p<sub>i</sub> | Recovery<br>median (IQR) | p<sub>R</sub> |
| PLACEBO<br>(N = 21) | 39.0<br>(33.0 – 41.0) | 9.10<br>(6.83 - 11.52) | **3.73×10<sup>-8</sup>** | 2.31<br>(1.19 - 4.34) | 0.318 |
| ESTRADIOL<br>(N = 15) | 38.0<br>(29.5 – 43.5) | -0.23<br>(-2.20 - 2.98) |  | 1.31<br>(-0.81 - 2.88) |  |

p values describe statistical significance of difference between estradiol and placebo group after intervention (p<sub>i</sub>) and recovery (p<sub>R</sub>)

**Figure 2.**
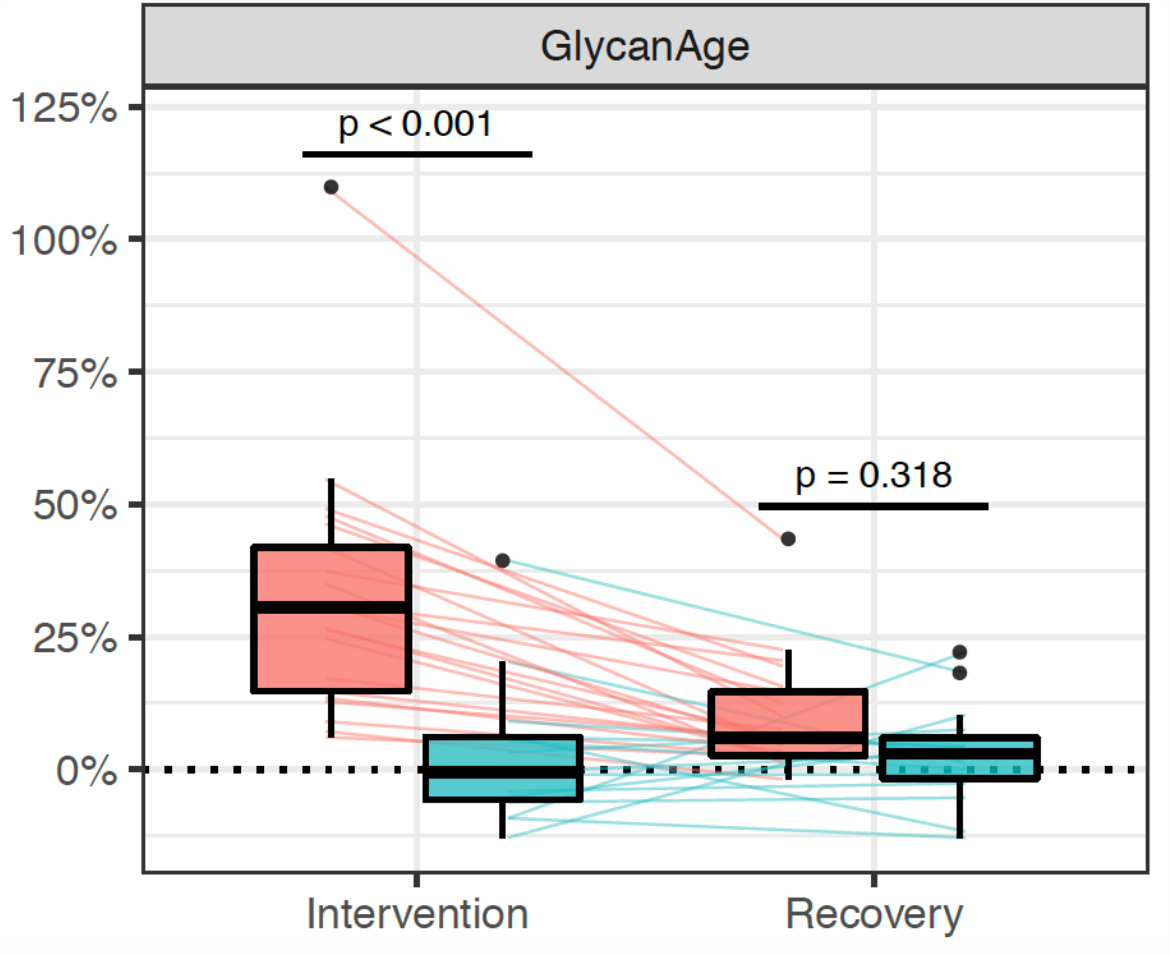
Distribution of changes in glycan age in 36 women undergoing gonadal hormone suppression for 6 months. Statistically significant increase in glycan age was observed in the placebo group (n=25, red rectangle), while supplementation with estradiol prevented this change (n = 15, blue rectangle)

**Figure 3.**
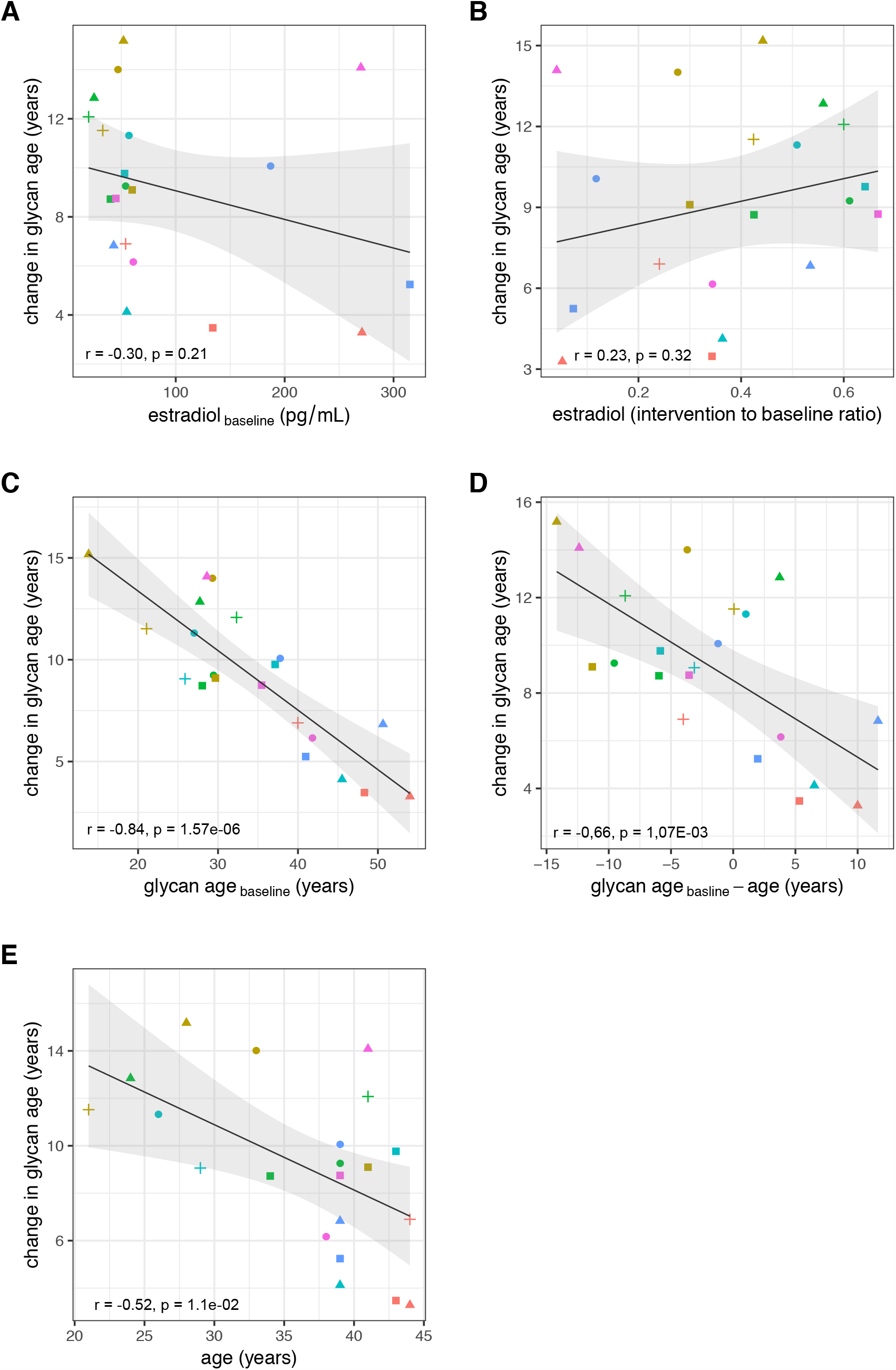
Correlations between the change in glycan age caused by gonadal hormone suppression and baseline estradiol concentration (A), change in estradiol concentration (b), glycan age (C), change in glycan age (D) and age.

## Discussion

One of the key requirements for an aging biomarker is that this it is responsive to interventions that are beneficially affecting the biology of ageing, but convincing evidence of this is still missing for any ageing biomarker [12]. In this study, we show that the removal of gonadal hormones results in a rapid increase of glycan age, which can be completely prevented by supplementation with estrogen hormone replacement, a therapy proven to be of benefit to some perimenopausal women [13]. Glycan age is a measure of biological age that is based on the analysis of IgG glycosylation [14]. Glycans attached to IgG are functionally important because they regulate inflammation at multiple levels [15,16] and are considered to be one of the important drivers of inflammaging [17]. Large studies of adult human populations indicated that IgG glycans without galactose and sialic acid that are the main component of the increased glycan age increase with the onset of menopause [1], while in girls they decrease with the onset of puberty [8,18]. This indicated that estrogen may be relevant, but since many things change during puberty and menopause, the change in glycan age could not have been directly attributed to the change in estrogen concentration. In our study suppression of gonadal hormones in premenopausal women resulted in considerable (median increase = 9.1 glycan age years) increase in glycan age that was statistically highly significant. Moreover, the change was observed in all study participants that received placebo instead of estradiol supplementation. At the same time, the supplementation with estradiol was sufficient to completely prevent the increase in biological age.

The extent of changes in both hormone levels (Table 1) and glycan age (Figure 2) varied considerably between individual participants. Aiming to determine what contributed to the extent of change within each individual we compared correlations of changes in glycan age levels with basal hormone levels, changes in hormone levels, basal glycan age level and the difference between glycan age and chronological age. We did not find a statistically significant correlation between the change in glycan age and the baseline serum estrogen concentration or the change in serum estrogen concentration after the intervention. However, both basal glycan age and the difference between chronological and glycan age were strongly negatively correlated with the change in glycan age. This suggests that despite being evidently important, estrogen is only one of the factors that define the glycan age of an individual.

Despite extensive research, progress in the development of biomarkers that could reliably quantify inter-individual differences in aging is still limited [19]. One of the important elements that is still missing is the ability to change the biomarker with lifestyle changes or pharmacological interventions. Recently a modest improvement in epigenetic age was reported in a small group of individuals undertaking quite radical pharmacological intervention [20] and glycan age was shown to slightly improve by exercise [21]. However, all these changes were modest compared to the effects of the suppression of gonadal hormones, which more than doubled glycan age in some of the participants. Supplementation with estradiol was sufficient to completely abolish this effect. It is intriguing to speculate that hormone supplementation could also prevent the increase of glycan age that occurs around menopause, but this still needs to be investigated. Furthermore, since IgG glycosylation is a functionally relevant modification that regulates the immune system, this discovery opens the option to look for downstream pathways that may be a more specific target for therapy than broadly acting estrogen.

## Methods

### Institutional approval

This study was conducted at the University of Colorado Anschutz Medical Campus (CU-AMC). Procedures followed were in accordance with the ethical standards of and approved by the Colorado Multiple Institutional Review Board (COMIRB) and the Scientific Advisory and Review Committee at the University of Colorado Anschutz Medical Campus (CU-AMC). The study was registered on ClinicalTrials.gov (NCT00687739) on May 28, 2008.

### Participants and screening procedures

Participants were healthy eumenorrheic premenopausal women (n = 36). In accordance with the Declaration of Helsinki, volunteers provided written informed consent to participate, with the knowledge that the risks of the study included menopause-like effects (e.g., weight gain, bone loss, menopausal symptoms). Main inclusion criteria were age (25 to 49 y) and normal menstrual cycle function (no missed cycles in previous year, cycle length 28±5 d and confirmation of ovulatory status (ClearPlan Easy, Unipath Diagnostics, Waltham, MA)). Exclusion criteria were pregnancy or lactation, use of hormonal contraception, oral glucocorticoids, or diabetes medications, smoking, or body mass index (BMI) >39 kg/m^2^. Volunteers underwent screening procedures, as described previously [11].

### Experimental design and study procedures

The parental trial was a randomized, double-blinded, placebo-controlled trial to determine the effects of estradiol (E_2_) deficiency on body composition and energy expenditure, bone mineral density, components of energy expenditure and physical activity in premenopausal women [11,22]. In Short, all participants underwent suppression of ovarian sex hormones with gonadotropin releasing hormone agonist therapy (GnRH_AG_, leuprolide acetate 3.75 mg, Lupron; TAP Pharmaceutical Products, Inc; Lake Forest, IL) in the form of monthly intramuscular injections. A single injection of leuprolide acetate produces an initial stimulation (for 1 to 3 wk) followed by a prolonged suppression of pituitary gonadotropins FSH and LH, while repeated monthly dosing suppresses ovarian hormone secretion [23]. The absence of pregnancy was confirmed by a urine pregnancy test before each dosing. After screening procedures were completed, eligible volunteers underwent baseline testing during the early folicular phase (days 2 to 6 after onset of menses) of the menstrual cycle. At the beginning of the next menstrual cycle participants began with 5-month GnRH_AG_ therapy to chronically suppress ovarian function. Participants were randomized to receive either transdermal E_2_ 0.075 mg/d (Bayer HealthCare Pharmaceuticals, Berkeley, CA) or placebo patches (GnRHAG+E2, n=15; GnRH_AG_+PL, n=21). In order to reduce the risk of endometrial hyperplasia, but in the same time minimizing the exposure to progesterone, women randomized to E2 received medroxyprogesterone acetate (5mg/d, as a pill) for 12 days every other month (end of month 2 and 4, and after completion of follow-up testing). During these monthly visits, participants were under supervision of the research nurse practitioner. Participants were asked to report changes in use of medications or health (e.g., doctor visits, hospitalizations), as well as any study-related problems/concerns over the past 4 weeks. The E_2_ regimen was expected to maintain serum E_2_concentrations in the mid-to-late follicular phase range (100 to 150 pg/mL).

### Sample collection

Blood samples for sex hormones and glycans were collected in three timepoints: during baseline testing (T1), during week 20 of the hormonal intervention (T2), and at the spontaneous recovery of the normal menstrual cycle function (T3). A single sample (∼5 mL) was obtained in the morning (∼8 AM), after an overnight fast (at least 10 hours). Baseline samples were obtained immediately before the first GnRH_AG_ injection. Serum was separated from each collected sample upon blood withdrawal and stored at −80°C until analysis.

### Sex hormones

Collected sera were analyzed for numerous sex hormones. Estrone (E1), estradiol E2 and progesterone (P) were determined by radioimmunoassay (RIA, Diagnostic Systems Lab, Webster, TX). Total testosterone (T) was analyzed by chemiluminescence immunoassay (Beckman Coulter, Inc. Fillerton, CA) and sex hormone-binding globulin (SHBG) by immunoradiometric assay (Diagnostic Systems Laboratory).

### N-glycosylation of immunoglobulin G

The whole procedure was performed according to already published protocol [24]. In short, IgG was isolated from sera samples by affinity chromatography using 96-well Protein G plate (BIA Separations, Slovenia). The isolated IgG was denaturated with the addition of SDS (Invitrogen, USA) and incubation at 65°C. The excess of SDS was neutralized by the addition of Igepal-CA630 (Sigma-Aldrich, USA) and N-glycans were released with the addition of PNGase F (Promega, USA) in PBS buffer followed by overnight incubation at 37°C. The released glycans were fluorescently labelled with 2-AB (Merck, Germany). Free label and reducing agent were removed from the samples by using hydrophilic interaction liquid chromatography solid phase extraction (HILIC-SPE). IgG N-glycans were eluted with ultrapure water and stored at −20°C until use. Fluorescently labelled N-glycans were separated using HILIC on an Acquity UPLC H Class Instrument (Waters, USA) that consists of sample manager, quaternary solvent manager and fluorescence (FLR) detector. The instrument was under the control of Empower 3 software, build 3471 (Waters, USA). Labelled N-glycans were separated on an amide ACQUITY UPLC® Glycan BEH chromatography column (Waters, USA), 100 × 2.1 mm i.d., 1.7 μm BEH particles, with 100 mM ammonium formate, pH 4.4, as solvent A and 100% ACN as solvent B. Samples were kept at 10°C before injection, and separation was performed at 60°C. The separation method used a linear gradient of 25-38% solvent A at a flow rate of 0.40 mL/min in a 27 min analytical run. Fluorescently labelled N-glycans were detected by FLR detector with excitation and emission wavelengths set at 250 and 428 nm. Data processing included an automatic integration algorithm that was manually corrected to maintain the same intervals of integration for all the samples. IgG N-glycan samples were all separated into 24 peaks and the relative amount of glycans in each chromatographic peak was expressed as percentage of total integrated area (% Area).

### Statistical analysis

Area under chromatogram peaks was normalized to total chromatogram area, then each glycan peak was logit transformed and batch corrected using ComBat method (R package ‘sva’)[25]. Data were back transformed, and derived glycan traits were calculated as a sum or ratio of selected directly measured glycan peaks based on particular glycosylation features (i.e. sialylation or fucosylation). GlycanAge was calculated according to Krištić *et al* [1]: age model coefficients were trained using 1116 females (18 – 98 years old) from The Croatian National Biobank “10 001 Dalmatians” [26]. IgG N-glycome from the biobank was measured in the same laboratory and prepared in the same way as estrogen-study data. Strength of the associations were estimated using Pearson’s correlation coefficient. All statistical analyses were performed in R programming software (version 3.6.3) [27].

## CONFLICT OF INTEREST STATEMENT

G. Lauc is the founder and owner of Genos Ltd, a private research organization that specializes in high-throughput glycomic analyses and has several patents in this field. J. Jurić, F. Vučković and M. Pezer are employees of Genos Ltd. G. Lauc is also co-founder of GlycanAge Ltd and is listed as an inventor on patents EP3011335B1 and US9910046B2 that protect the use of glycan age to predict biological age. All other authors declare no conflicts of interest.

## Data Availability

All data is available upon reasonable request

## ACKNOWLEDGEMENTS

Glycosylation analysis was performed in Genos Glycoscience Research Laboratory and partly supported by the European Structural and Investment Funds IRI grant (#KK.01.2.1.01.0003), Centre of Competence in Molecular Diagnostics (#KK.01.2.2.03.0006) and Croatian National Centre of Research Excellence in Personalized Healthcare grant (#KK.01.1.1.01.0010). PAN was supported by the US National Institute of Arthritis and Musculoskeletal and Skin Diseases grant P30AR070253. We acknowledge the members of our research groups who carried out the day-to-day activities for the project. Finally, we want to thank the women who volunteered to participate in the study for their time and efforts.

## Notes

### Clinical Trial

The intervention study was registered on ClinicalTrials.gov (NCT00687739) on May 28, 2008. Glycan analysis performed for this study was not available in 2008 and was not originally planned. It was performedd in 2020 in leftover plasma samples from the trial.

